# Resistance-minimizing strategies for introducing a novel antibiotic for gonorrhea treatment: a mathematical modeling study

**DOI:** 10.1101/2023.02.14.23285710

**Authors:** E Reichert, R Yaesoubi, MM Rönn, TL Gift, JA Salomon, YH Grad

**Affiliations:** Department of Immunology and Infectious Diseases, Harvard T. H. Chan School of Public Health, Boston, Massachusetts, USA; Department of Health Policy and Management, Yale School of Public Health, New Haven, Connecticut, USA; Department of Global Health and Population, Harvard T.H. Chan School of Public Health, Boston, MA, USA; Division of STD Prevention, Centers for Disease Control and Prevention, Atlanta, Georgia, USA; Department of Health Policy, Stanford University School of Medicine, Stanford, CA, USA

**Keywords:** antibiotic resistance, gonorrhea, mathematical modeling, antibiotic introduction

## Abstract

**Background:** Gonorrhea is a highly prevalent sexually transmitted infection and an urgent public health concern due to increasing antibiotic resistance. Only ceftriaxone remains as the recommended treatment in the U.S. The prospect of approval of new anti-gonococcal antibiotics raises the question of how to deploy a new drug to maximize its clinically useful lifespan.

**Methods:** We used a compartmental model of gonorrhea transmission in the U.S. population of men who have sex with men to compare strategies for introducing a new antibiotic for gonorrhea treatment. The strategies tested included holding the new antibiotic in reserve until the current therapy reached a threshold prevalence of resistance; using either drug, considering immediate and gradual introduction of the new drug; and combination therapy. The primary outcome of interest was the time until 5% prevalence of resistance to both the novel drug and to the current first-line drug (ceftriaxone).

**Findings:** The reserve strategy was consistently inferior for mitigating antibiotic resistance under the parameter space explored. The reserve strategy was increasingly outperformed by the other strategies as the probability of *de novo* resistance emergence decreased and as the fitness costs associated with resistance increased. Combination therapy tended to prolong the development of antibiotic resistance and minimize the number of annual gonococcal infections.

**Interpretation:** Our study argues for rapid introduction of new anti-gonococcal antibiotics, recognizing that the feasibility of each strategy must incorporate cost, safety, and other practical concerns. The analyses should be revisited once robust estimates of key parameters–likelihood of emergence of resistance and fitness costs of resistance for the new antibiotic–are available.

**Funding:** U.S. Centers for Disease Control and Prevention (CDC), National Institute of Allergy and Infectious Diseases

## Introduction

Increasing antibiotic resistance in *Neisseria gonorrhoeae* poses an urgent clinical and public health threat. Only one antibiotic – ceftriaxone, an extended spectrum cephalosporin – remains recommended by U.S. CDC guidelines for empirical treatment of gonorrhea(1), underscoring the need to develop new antibiotics to treat this highly prevalent sexually transmitted infection (STI). Two promising, first-in-class candidates in phase III clinical trials are zoliflodacin (Entasis Therapeutics) and gepotidacin (GlaxoSmithKline). In phase II trials, both demonstrated cure rates of 96% for urogenital infection; cure rates for pharyngeal infection, historically harder to cure, were lower(2,3). Given the limited tools remaining for gonorrhea treatment, it is imperative to deploy new antibiotics in a way that prolongs their clinical effectiveness.

The question of how best to utilize multiple effective antibiotics with the aim of minimizing population-level resistance has been explored in mathematical modeling studies considering a range of strategies: I) random allocation of multiple drugs, II) combination therapy, III) holding in reserve a second-line drug until prevalence of resistance to the first-line drug reaches a defined threshold, IV) cycling treatment, and V) targeted (directed) treatment, where a second-line drug is given only to those non-responsive to the first(4–11). One pervasive finding across these studies is that antibiotic cycling is inferior for mitigating antibiotic resistance; however, among alternative approaches the optimal strategy varies by study setting and pathogen of interest. While one study focused specifically on gonorrhea(11), none explored how to optimally introduce a new antibiotic.

Historically, U.S. guidelines for gonorrhea treatment have followed several strategies. Multiple options enabled providers to choose antibiotics during the 1990s. Increasing resistance then led to combination therapy (ceftriaxone plus azithromycin), and recently, rising azithromycin resistance prompted a return to single drug treatment with ceftriaxone(12). However, the question of how to best allocate multiple antibiotics will once again become relevant if zoliflodacin, gepotidacin, or another candidate gains FDA approval. Therefore, this study aims to compare the impact of introduction strategies for a novel antibiotic on resistance of *N. gonorrhoeae* among the U.S. population of men who have sex with men (MSM), using a mathematical model of gonorrhea transmission. We focused on MSM as approximately one-third of gonorrhea cases in the U.S. occur among MSM(13), and due the historic appearance of resistance to antibiotics like ciprofloxacin within this population(14), gonorrhea transmission models characterizing U.S. MSM have been most well-researched(15–17).

## Methods

### Model Overview

We modified the susceptible-infectious-susceptible (SIS) deterministic compartmental gonorrhea transmission model described in Tuite et al. 2017(17) (Figure 1 and Technical Supplement). In brief, the model consists of an MSM population stratified into 3 sexual activity groups characterized by annual rates of partner change. Sexual activity group was fixed for each individual, but individuals of different sexual activity groups interact with mixing parameter ε. Individuals age into and out of the sexually active population at rate ρ, contributing for 20 years on average. Infected individuals can recover spontaneously or through antibiotic treatment. For those that seek treatment, one can receive drug A (ceftriaxone-like antibiotic) and/or drug B (new antibiotic). Infections are stratified by symptomatic (Y) versus asymptomatic status and by resistance profile, where each infection can be resistant to drug A, drug B, neither, or both. Note that we use the term ‘individual’ only to help conceptualize the interaction and movement of this population between model compartments.

**Figure 1.**
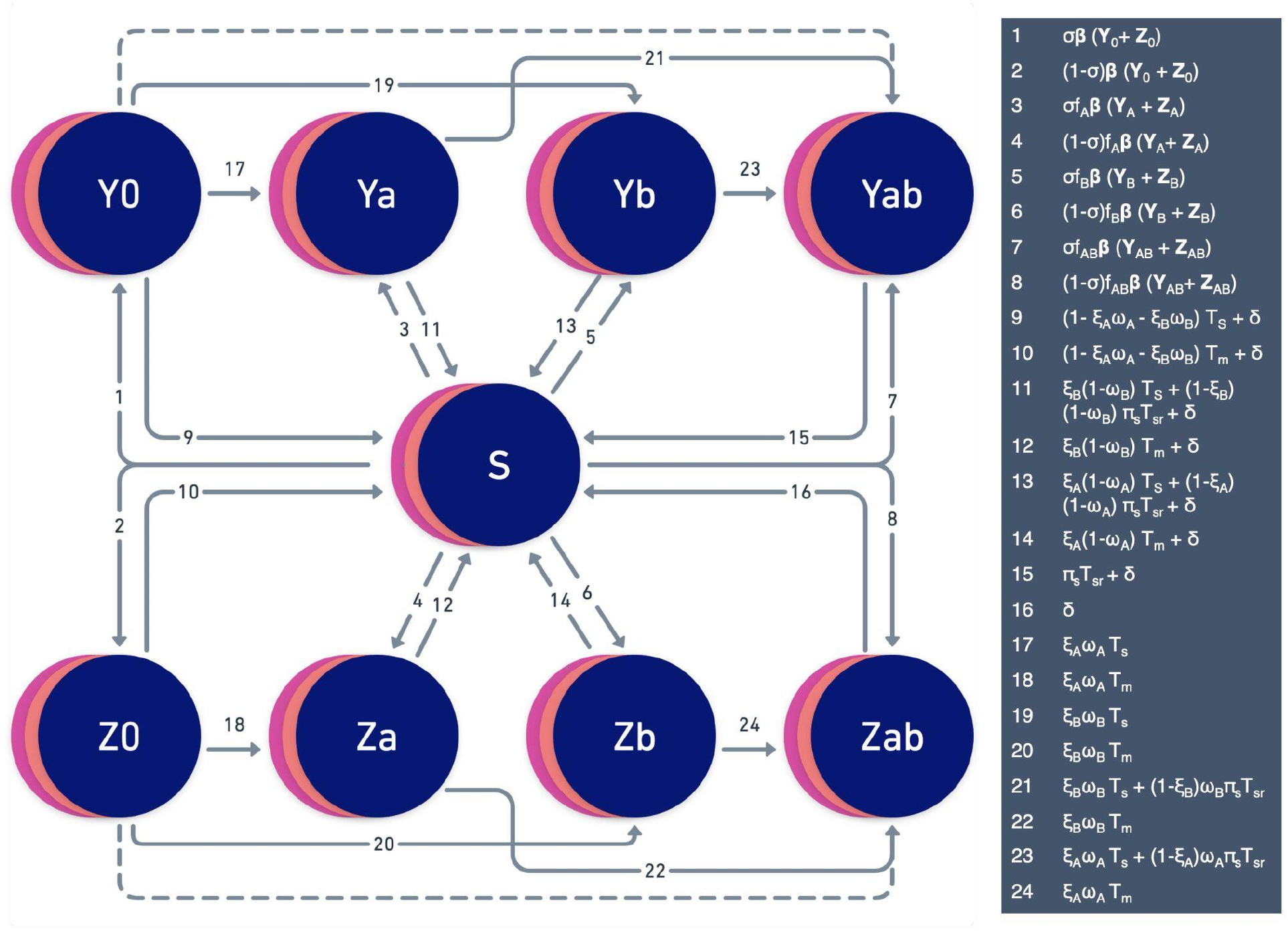
Schematic of gonorrhea transmission model. Abbreviations: S = Susceptible, Y = Symptomatic Infection, Z = Asymptomatic Infection. Infections are further stratified by resistance profile, where 0 = susceptible, a = resistant to drug A, b = resistant to drug B, and ab = resistant to both drugs. Overlapping discs for all compartments represent the model’s stratification into three sexual activity groups: low, intermediate, and high. Arrows depict rates between compartments and would be multiplied by the compartment from which they flow to generate the model’s set of differential equations (see Technical Supplement). Dotted arrows represent transitions only possible with combination therapy. Individuals can also enter and exit the population at rate ρ (arrows not shown). Rates shown here apply to random 50-50 allocation, reserve, and gradual switch strategies; rates for combination treatment vary slightly and can be found in the Technical Supplement. Definitions of all parameters used in rate equations can be found in Table 1.

### Model Parameterization

Model parameters were determined from the literature or model calibration using maximum likelihood estimation (MLE; Table 1). For parameters relating to gonorrhea’s natural history and the population’s sexual behavior, initial values were assigned from parameter estimates in Tuite et al. 2017(17). We then calibrated our model to a beta distribution with mean 3.0% gonorrhea prevalence (variance 1.47 × 10^−5^) at equilibrium, based on recent estimates in MSM(13,18,19). Model simulations for calibration were run for a drug A-only model over two years and parameterized using the R package bblme, which chose a set of parameter values that optimized the likelihood of the simulated gonorrhea prevalence, assuming a Beta(α = 59.36, β = 1919) prevalence distribution (20).

**Table 1.** Model parameters

|  | Description | Value | Source(s) |
| --- | --- | --- | --- |
| *Model Calibration Target* |  |  |  |
|  | Gonorrhea prevalence at start (Calibration target mean) | 3.0% |  |
| *Model Population, Sexual Behavior parameters* |  |  |  |
| N | Population size | $10^6$ | Assumption |
| n | Relative size of sexual activity groups |  | Tuite et al. 2017(17) |
| | Low | $n_1 = 0.3$ | |
| | Intermediate | $n_2 = 0.6$ | |
| | High | $n_3 = 0.1$ | |
| $\theta$ | Rate of partner change per sexual activity group (per year) | | Model fitting; Tuite et al. 2017 |
| | Low | $\theta_1 = 1 \times 1.22$ | |
| | Intermediate | $\theta_2 = 5 \times 1.22$ | |
| | High | $\theta_3 = 20 \times 1.22$ | |
| $\epsilon$ | Mixing parameter | 0.24 | Model fitting; Tuite et al. 2017 |
|  | Proportion of cases drug A (ceftriaxone-like) resistant at start | 0.0001 | CDC GISP 2020(12), Tuite et al. 2017 |
|  | Proportion of cases drug B (new drug) resistant at start | 0 | Assumption |
| $\rho$ | Model entry/exit rate (per year) | 1/20 | Tuite et al. 2017 |

| *Gonorrhea Natural History parameters* |  |  |  |
| --- | --- | --- | --- |
| $\sigma$ | Proportion of **incident** infections that are symptomatic | 0.60 | Model fitting; Tuite et al. 2017 |
| $b$ | Transmission probability per partnership | 0.46 | Model fitting; Tuite et al. 2017, Fingerhuth et al. 2016(15), Tuite et al. 2018(22) |
| $\delta$ | Natural recovery rate from infection (per year) | 1/0.462 | Model fitting; Tuite et al. 2017, Vegvari et al. 2020(23) |
| *Treatment parameters* |  |  |  |
| $T_s$ | Treatment rate if initial treatment success, symptomatic infection (per year) | 1/0.031 | Model fitting; Tuite et al. 2017, Tuite et al. 2018 |
| $T_{sr}$ | Treatment rate if initial treatment failure (requiring retreatment), symptomatic infection (per year) | $T_s/3$ | Model fitting; Tuite et al. 2017 |
| $T_m$ | Screening rate, asymptomatic infection (per year) | 0.40 | Model fitting; Tuite et al. 2017, Hui et al. 2013(24), Tuite et al. 2018 |
| $\xi$ | Probability of receiving drug upon initial treatment | | Assumption |
| | Drug A ($\xi_A$) | Strategy dependent | |
| | Drug B ($\xi_B$) | Strategy dependent | |
| $\omega$ | Probability of emergence of resistance upon treatment | | |
| | Drug A | $\omega_A = 10^{-8}$ | Tuite et al. 2017, Vegvari et al. 2020 |
| | Drug B | $\omega_B = 10^{-4} (10^{-10}-10^{-2})$ | Assumption (range) |
| | Drugs A and B | $\omega_A * \omega_B$ | Assumption |
| $f$ | Relative fitness of resistant bacteria, compared to susceptible | | |
| | A resistant | $f_A = 0.98$ | Tuite et al. 2017 |
| | B resistant | $f_B = 0.95 (0.80-1)$ | Assumption (range) |
| | Dual resistance | $f_{AB} = f_A * f_B$ | Assumption |
| $\pi_s$ | Proportion receiving retreatment if initial treatment failure, symptomatic infection | 0.90 | Tuite et al. 2017 |

Antimicrobial susceptibility surveillance data from the Gonococcal Isolate Surveillance Project (GISP) informed a conservative estimate of the percentage of infections resistant to the ceftriaxone-like drug (0.01%) at time 0(12). Relevant properties of ceftriaxone, including the probability of *de novo* resistance (ω_A_) and the fitness cost associated with resistance (1-f_A_), were inferred from prior literature(17). We used ‘drug A’ to emphasize that we are modeling a ceftriaxone-like drug, as its parameters are only informed estimates of ceftriaxone’s properties. All gonococcal infections were presumed susceptible to the novel antibiotic, drug B, at time 0. Baseline properties of drug B, including the probability of emergence of *de novo* resistance upon treatment (ω_B_) and the associated fitness cost (1-f_B_), were partially informed by the limited data we have from phase II clinical trials for gepotidacin and zoliflodacin. The baseline probability of *de novo* resistance for drug B was greater than for drug A given gepotidacin’s phase II results suggesting failure upon treatment is more common than with ceftriaxone(3). The baseline relative fitness of strains resistant to drug B was assumed lower than for drug A in keeping with reports that GyrB mutations associated with zoliflodacin resistance incur a substantial fitness cost(21). However, absolute parameter estimates were assumed and further explored in the sensitivity analysis described below. We assumed resistance mechanisms to drugs A and B were independent.

### Antibiotic Introduction Strategies

We compared four different strategies for the deployment of drug B into the population, described below:

*1) Random 50-50 Allocation*. Each treatment-seeking infected individual has a 50% probability of receiving either drug A or drug B, effective at time 0.
*2) Combination Therapy*. Each treatment-seeking infected individual receives both drug A and drug B, effective at time 0.
*3) Reserve Strategy*. Each treatment-seeking infected individual receives drug A until a 5% prevalence of resistance is reached in the population, at which point drug B is introduced with a sigmoid growth function until it is used for 100% of presenting cases.
*4) Gradual Switch*. Each treatment-seeking infected individual receives drug A at time 0, but drug B is gradually introduced into the population until complete random 50-50 allocation is reached within 9.5 years (midpoint = 4 years).

A threshold of 5% resistance prevalence was used to trigger the switch from drug A to B under the reserve strategy, as this represents the World Health Organization’s threshold for changing treatment recommendations(25). We incorporated a gradual switch strategy into our analysis to depict a more realistic introduction scenario for a first-in-class antibiotic, assuming factors such as clinician hesitancy, drug supply and distribution may delay uptake. For both the reserve and gradual switch strategies, the probabilities of receiving drugs A and B (*ξ*_A_, *ξ*_B_) for treatment-seeking individuals are not constant but updated throughout the model using sigmoid functions (Supp. Figure 1).

Of note, the above scenarios describe only the allocation of one’s initial course of treatment. Symptomatic individuals prescribed a drug ineffective against their infection can seek retreatment at rate T_sr_ with probability π_s_, at which point they will receive the alternative antibiotic, incorporating an element of targeted treatment into all approaches. An exception is assumed for symptomatic individuals with dual resistant infection who can also seek retreatment at rate T_sr_ with probability π_s_; for these cases, we assume retreatment with a last-resort antibiotic external to our model for which we assume complete efficacy and do not monitor resistance trends.

### Model Implementation

The base model was initialized with the equilibrium prevalence of gonorrhea achieved via calibration (3.0%) and simulated over 40 years using the R package deSolve(26). We compared the time until each of drugs A and B lost clinical effectiveness for empiric use, defined by reaching a 5% prevalence of resistance among gonococcal infections. We define this time to loss outcome (T_L_) by the maximum of i) the time to 5% resistance to drug A and ii) the time to 5% resistance to drug B. Other secondary outcomes explored include the time to a 1% prevalence of resistance threshold for each drug, as well as population-level prevalence, incidence, and cumulative number of incident gonococcal infections.

Since baseline parameters describing the properties of the novel drug B were assumed, we performed sensitivity analyses to compare strategies across a large parameter space for both i) the probability of resistance emergence upon treatment with drug B (ω_B_), and ii) the relative fitness associated with resistance to drug B (f_B_). Finally, since parameters are not known with certainty for drug A (ceftriaxone-like antibiotic), we reran the sensitivity analysis exploring drug B’s properties under two alternate scenarios for drug A: 1) fitness cost for drug A resistant strains (1-f_A_) increased to 0.10, and 2) probability of *de novo* resistance for drug A (ω_A_) increased to 10^−4^.

All code needed to run the model and produce numeric output and figures is available at https://github.com/emreichert13/gc-antibioticintro.

## Results

The transmission model projected that continued monotherapy with drug A (ceftriaxone-like antibiotic) for gonorrhea treatment would result in a 5% prevalence of resistance warranting new treatment guidelines within 6.5 years in our model population (Table 2; Supp. Fig. 2). Assuming introduction of the novel antibiotic B, with baseline parameters for its rate of *de novo* resistance emergence and associated fitness cost, time to loss (i.e., a 5% resistance threshold) of drug A ranged from 6.5 to 19.9 years depending on the introduction strategy, whereas drug B’s clinical effectiveness ranged between 13.9 to 19.9 years (Table 2; Supp. Fig 2).

**Table 2.** Time to predefined 1% and 5% resistance thresholds by antibiotic introduction strategy under baseline model conditions. T_L_ = time in years until both drugs A and B hit their 5% resistance thresholds, warranting new treatment recommendations. The range is bounded by the absolute minimum and maximum annual result.

| | Time to 1% Resistance Threshold (years) | | | Time to 5% Resistance Threshold (years) | | | End of Drugs' Lifespan ($T_L$) | Prevalence of Gonorrhea at $T_L$ | Average Number of Incident Infections per Year, $t < T_L$ (Range) |
| --- | --- | --- | --- | --- | --- | --- | --- | --- | --- |
|  | Drug A | Drug B | Dual | Drug A | Drug B | Dual |  |  |  |
| 1. Random 50-50 Allocation | 15.0 | 16.3 | 17.2 | 18.7 | 19.2 | 19.5 | 19.2 | 3.16% | 179,023<br>(178,005 - 182,998) |
| 2. Combination therapy | 17.7 | 17.7 | 17.7 | 19.9 | 19.9 | 19.9 | 19.9 | 3.14% | 178,641<br>(177,998 - 181,731) |
| 3. Reserve strategy | 4.7 | 11.8 | 14.6 | 6.5 | 13.9 | 17.9 | 13.9 | 3.13% | 180,084<br>(178,011 - 184,405) |
| 4. Gradual Switch to 50-50 | 7.9 | 15.6 | 15.8 | 13.6 | 18.2 | 18.2 | 18.2 | 3.36% | 180,983<br>(178,011 - 191,703) |
| *Monotherapy,*<br>*Drug A* | 4.7 | NA | NA | 6.5 | NA | NA | 6.5 | 3.15% | 179,220<br>(178,011 - 182,179) |

**Figure 2.**
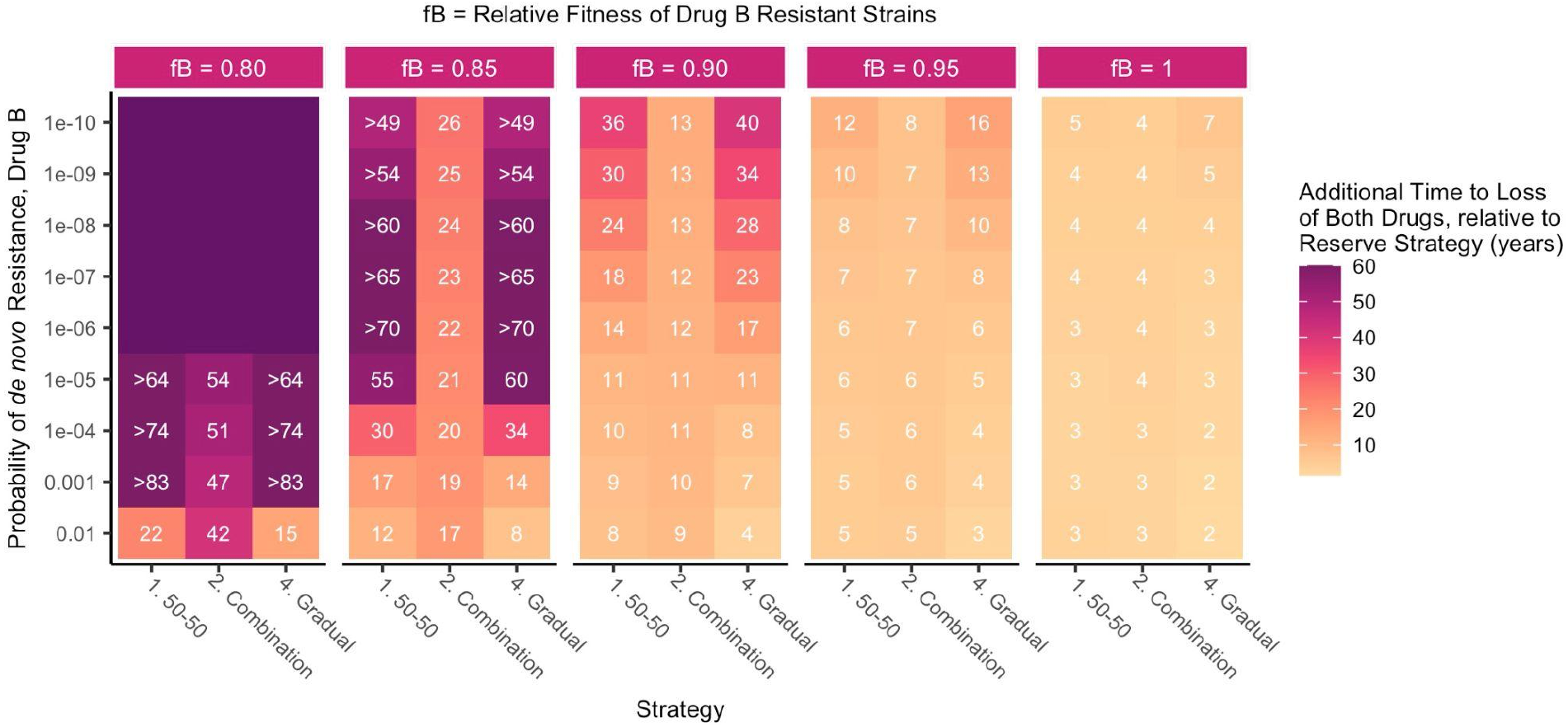
Additional time (in years) to loss of each of antibiotics A and B by strategy relative to that of the reserve strategy (T_L_|strategy - T_L_|reserve), based on properties of a new antibiotic B. Antibiotic A = ceftriaxone-like, Antibiotic B = new antibiotic. Strategies (x-axis) were compared over a range of plausible parameter values for the probability of emergence of resistance upon treatment with drug B (ω_B_; y-axis) and the relative fitness of drug B resistant strains (f_B_; vertical facets). These properties were held constant at 0.98 and 10^−8^ respectively for drug A. If the lifespan of these drugs extended beyond the 100 years over which the model was run, that strategy’s results are shown either in relative terms for comparison or with an unlabeled dark purple tile, if no strategies on the x-axis had a defined T_L_ under that parameter set for drug B. Abbreviations: fB = f_B_ = fitness of drug B resistant strains relative to susceptible bacteria, T_L_ = time in years until both drugs A and B hit their 5% resistance thresholds, warranting new treatment recommendations.

Combination therapy maximized the time until 5% resistance was met for both drugs (T_L_ = 19.9 years); random 50-50 allocation was a close alternative, reducing the time until the drugs were lost by less than one year to T_L_ = 19.2 years (Table 2; Supp. Fig. 2). In contrast, the reserve strategy led to a faster buildup of infections resistant to drug A; a 5% prevalence of resistance to drug A was present within 6.5 years. This facilitated an earlier emergence of dual resistance and resulted in the shortest combined lifespan of the drugs for the reserve strategy (T_L_ = 13.9 years) relative to alternative approaches.

Combination therapy also minimized the average annual number of incident gonococcal infections up until T_L_ (Table 2). Except for the reserve strategy, every strategy involving the introduction of drug B saw dual resistant strains rise to comprise >99% of infections after the treatments were lost given sufficient time and a lack of alternative antibiotics; this led to the model re-equilibrating at a gonorrhea prevalence approximately 3 times that at baseline (Supp. Fig. 2). Under the reserve strategy, since the overlap of drug A and drug B’s use was minimized, strains resistant to drug B-only took over following T_L_. However, if combination therapy was enacted after the loss of drug B at 13.9 years, dual resistant strains quickly rose to make up >99% of infections (Supp. Fig. 2). While the reserve strategy minimized time to loss T_L_, it in turn maximized the incidence of gonococcal infection over time (Supp. Figure 3).

**Figure 3.**
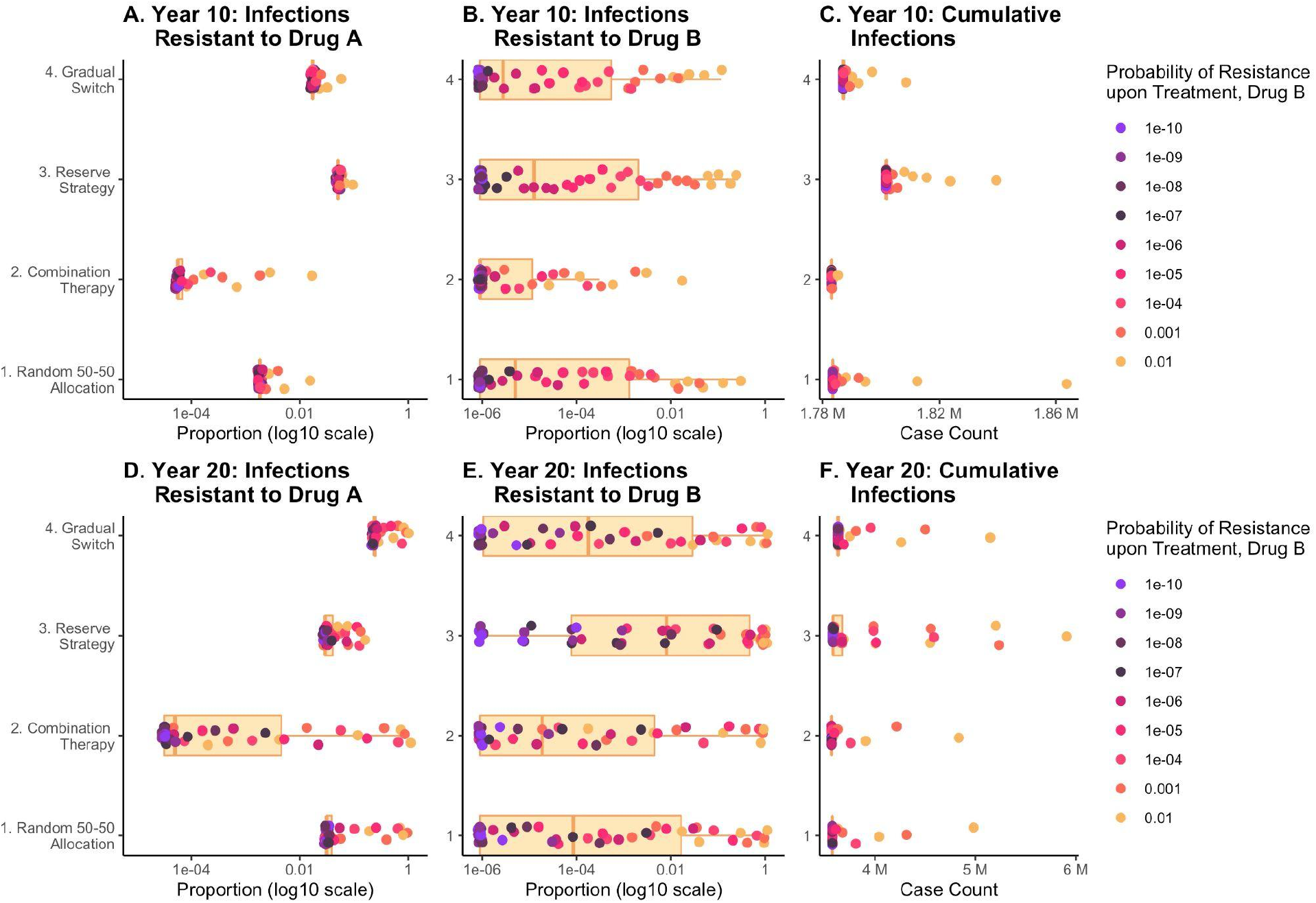
Distribution of antibiotic resistance and cumulative infections at Year 10 (top) and Year 20 (bottom), by introduction strategy. Each point represents the outcome from a different model run over one of 45 parameter sets for the relative fitness of bacteria resistant to drug B (f_B_ = 0.80-1) and the probability of resistance upon treatment (ω_B_ = 10^−10^ to 10^−2^). Points are colored by drug B’s probability of resistance upon treatment. Boxplots summarize the distribution of these outcomes across the 45 model runs by introduction strategy. These properties were held constant at f_A_ = 0.98 and ω_A_ =10^−8^ for drug A.

Model simulations were then run for 100 years over a larger, two-dimensional parameter space for the properties of drug B. The model run time was extended because some parameter sets increased the lifespan of available antibiotics >40 years for all strategies. The probability of emergence of resistance upon treatment with drug B (ω_B_) was varied from 10^−2^ to 10^−10^, and the fitness cost associated with resistance (1-f_B_) was varied from 0-0.20. Assuming baseline parameters for drug A, keeping the novel antibiotic B in reserve until drug A’s failure was not favorable under any scenario as it minimized the time until each of the drugs were lost for empiric use (T_L_; Figure 2). The combined lifespan of the drugs under the reserve strategy lagged behind alternative approaches by 2-3 years if resistance to drug B was likely to emerge (ω_B_ ≥ 10^−4^) and spread (fB = 1). Altering drug B’s characteristics to make the emergence and spread of resistance more unlikely resulted in the reserve strategy performing worse by greater margins compared to alternatives (Figure 2).

We then explored the distribution of the prevalence of drug A and drug B resistant infections, as well as the cumulative number of incident gonococcal infections, at two fixed points in time (Figure 3). For each introduction strategy, we explored 45 potential combinations of parameters for drug B, again varying the probability of *de novo* resistance (ω_B_ = 10^−10^-10^−2^) and relative fitness of resistant strains (f_B_ = 0.80-1). This allowed within- and between-strategy comparison of outcomes at 10 and 20 years. The median proportion of infections resistant to drug A ranged from 0.006 % (combination therapy) to 5.08% (reserve) at 10 years, and from 0.005%(combination therapy) to 23.4% (gradual switch) at 20 years.For drug B,the median proportion of resistant infections was ≤ 0.001% at 10 years for all strategies,but ranged from 0.002% (combination therapy) to 0.80% (reserve) at 20 years. The cumulative number of incident gonococcal infections ranged from a median of 1.78×10^6^ (combination therapy) to 1.80×10^6^ (reserve) cases at 10 years and from 3.57×10^6^ (combination) to 3.63×10^6^ (gradual switch) by year 20.

Finally, to account for uncertainty in the parameter estimates for ceftriaxone, we evaluated two scenarios: 1) fitness cost for drug A resistant strains (1-f_A_) increased to 0.10, and 2) probability of *de novo* resistance for drug A (ω_A_) increased to 10^−4^. In each case, we repeated the comparative analysis of the 45 different combinations of parameter values for drug B. While absolute estimates varied, the additional time to loss of each of drugs A and B (T_L_) relative to that of the reserve strategy remained positive across all introduction strategies and model runs (Supp. Fig. 4). The time by which other strategies extended the empiric use of drugs A and B relative to the reserve strategy generally increased in magnitude as both the probability of resistance emergence and the relative fitness of resistant strains decreased.

## Discussion

This study demonstrated that in a model of gonorrhea transmission in a population representative of U.S. MSM, among strategies to introduce a new antibiotic with the aim of slowing the spread of resistance, the strategy of reserving the novel antibiotic until substantial resistance has arisen to the current first-line drug is inferior to alternative strategies that introduce the novel antibiotic earlier. The impact of introducing the novel drug into the population once it becomes available varies depending on its likelihood of acquiring resistance and the fitness cost of resistance. As resistance to drug B becomes less likely to emerge and more costly, the reserve strategy is increasingly outperformed by alternative introduction strategies in terms of extending the clinical effectiveness of the available antibiotics.

Similar to findings of previous studies(4,6,9,10), no single introduction strategy for a new antibiotic targeting *N. gonorrhoeae* proves robustly optimal in the long term. Under baseline model assumptions, combination therapy maximized the clinical utility of the antibiotics and minimized the average number of annual infections. Across all parameter sets explored for drug B, combination therapy also minimized cumulative infections at 10 and 20 years on average. While the eventual emergence of dual resistant gonorrhea strains made combination therapy comparable to other strategies over longer time intervals, its benefits were seen most starkly in the short-term, as it delayed emergence of resistant gonococcal strains. As the probability of resistance upon drug B treatment decreased, other strategies became more favorable than combination therapy in maximizing the drugs’ combined lifespan (Figure 2). This indicates that if the probability of resistance acquisition is already very small, combination therapy loses some advantage.

Substantial between-strategy variation was found in the proportion of infections resistant to drug A at 10 and 20 years, despite drug A’s fixed parameters (Fig. 3A, 3D). This exhibits that the introduction strategy used for drug B has consequences on resistance trends to drug A.

Our analyses focused on the time to loss of the drugs (T_L_), as beyond this point neither drug would remain recommended for empiric gonorrhea treatment. Interpretation of results after T_L_ warrants caution. For example, the reserve strategy may initially appear attractive (Supp. Fig 2), as it precludes the takeover of dual-resistant strains. However, after loss of drug B at 13.9 years, combination therapy could be enacted to successfully treat cases resistant to drug B only, but only for a short time until dual resistance takes over. Short-term model outcomes (Figure 3) are also more relevant than long-term projections, as new tools for gonorrhea management and prevention (e.g., rapid antimicrobial susceptibility diagnostics and vaccines) in development may impact *N. gonorrhoeae*’s dynamics. Of note, the reserve strategy lengthened the projected time between the loss of drugs A and B; although not inherently clinically relevant, having this lag may be more palatable to medical providers than losing both drugs in rapid succession.

Other factors beyond those explored here inform how to best deploy a new antibiotic. The cost of a novel therapeutic is a major factor in determining its use, as are safety, tolerability, allergy, mode and duration of administration, and drug interaction concerns. This analysis can best be interpreted as an exploration of the ideal antibiotic introduction strategy for deterring resistance in *N. gonorrhoeae* assuming that these other practical concerns are not prohibitive (and keeping in mind key model limitations discussed below). For example, through exploring combination therapy, we assumed that this option is clinically safe and effective – that is, drugs A and B have compatible pharmacokinetics, no antagonistic effects, and using a therapeutic dose of both in combination is non-toxic. The feasibility and effectiveness of combination therapy relies on these assumptions. A disparity in the pharmacokinetics of ceftriaxone and azithromycin is one hypothesis for why combination therapy failed to deter resistance to azithromycin – azithromycin’s long elimination half-life as compared to ceftriaxone could leave hosts exposed to only a sub-inhibitory azithromycin concentration, an environment that would select for resistance given recurrent gonococcal infection(27).

We are further limited in that we ignore bystander selection, or selection for resistance caused by gonorrhea-infected individuals receiving antibiotic treatment for other indications. The relative importance of bystander compared to direct selection for resistance in *N. gonorrhoeae* is still unclear, and further research is needed to clarify its extent(27). Ceftriaxone is used to treat a variety of bacterial infections in inpatient settings; evidence that bystander selection is a major driver of *N. gonorrhoeae* resistance would support restricting use of a novel antibiotic to gonorrhea indications. We also do not consider the possible importation of drug resistant strains to this U.S. population from other countries with different treatment policies.

Many of the model parameters describing gonorrhea’s natural history and transmission are impossible to measure empirically, so we estimate these values with maximum likelihood estimation, an optimization procedure that here uses a single equilibrium prevalence target for model calibration. Prevalence estimates — of gonorrhea as well as ceftriaxone-resistant strains – are potentially underestimated due to missed asymptomatic infections. Further, estimating multiple parameters using a single prevalence estimate for calibration suggests that individual parameters would vary under alternative calibration targets or procedures. We also make the assumption that there is no resistance to the novel antibiotic B prior to its introduction, which may or may not hold. Amino acid mutations in GyrB associated with zoliflodacin resistance were found at a 0% prevalence in a large *N. gonorrhoeae* genome database(28), but mutations associated with gepotidacin resistance appeared in isolates resistant to tetracycline, ciprofloxacin, and penicillin(29), suggesting that gepotidacin resistance may be more easily acquired in a subset of the bacterial population. Absolute estimates of the time to 5% resistance prevalence for both drugs should therefore be interpreted with caution; results are best suited for relative interpretation, consistent with our aim of comparing the various introduction strategies.

It is also key to acknowledge that our model focuses specifically on the MSM population; however, trends in gonorrhea resistance in MSM in the U.S. have historically been forewarnings for resistance trends in the general population(13,14). Further, the model does not differentiate infections by anatomical site, which may be relevant given the drug B candidates’ variable efficacy in curing non-urogenital infection(2,3). We also assume uniform screening and treatment-seeking behavior across MSM by infection type (asymptomatic vs. symptomatic).

Overall, the steady accumulation of antibiotic resistance in *N. gonorrhoeae* and rising gonorrhea rates underscore the importance of optimizing all tools that prevent and treat infection. The results here suggest “reserve” strategies are least optimal and favor combination therapy, though it will be important to revisit these conclusions with updated estimates of the rates of emergence of resistance and fitness cost of resistance, as well as with the impact of additional interventions as they become available.

## Data Availability

All code needed to simulate the data and run analysis are available at https://github.com/emreichert13/gc-antibioticintro.

## Author Contributions

YG can be credited with study conceptualization, design, and supervision. ER developed the mathematical model, simulated modeled data, and conducted the analysis. ER and YG wrote the original draft. All authors participated in manuscript review and editing and were responsible for the final decision to submit for publication.

## Acknowledgements

We thank Ashleigh Tuite and colleagues for their deterministic compartmental gonorrhea transmission model in MSM(17) which was adapted for this research study.

## Ethical Approval

No ethical approval was required for this modeling study.

## Supplementary Material

**Supplementary Figure 1.**
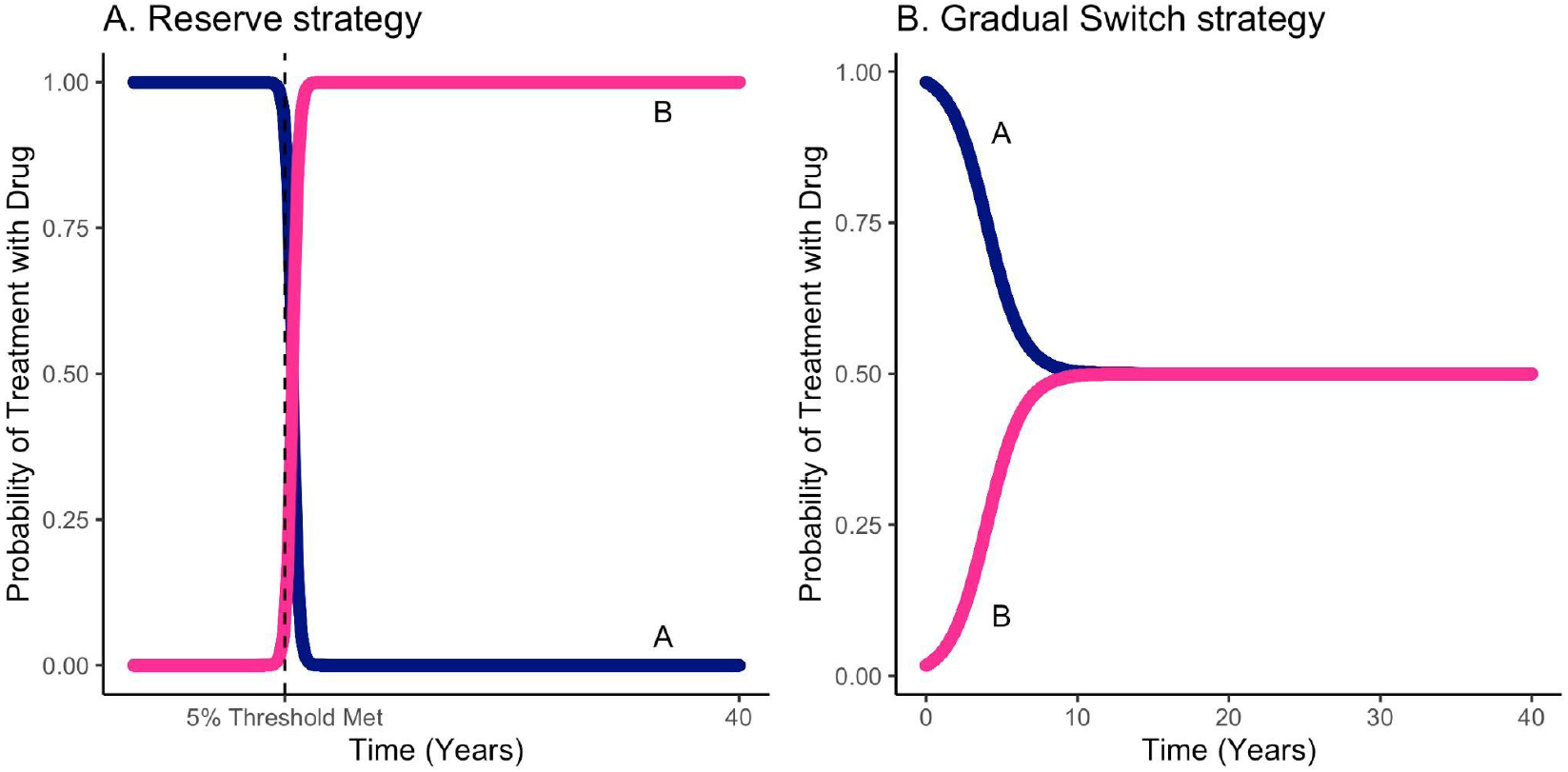
Sigmoid functions that define probability of treatment with drugs A and B (*ξ*_A_, *ξ*_B_) under the A) reserve and B) gradual switch strategies. A) The probability of receiving each drug is updated over time; a change is triggered at the time drug A reaches the 5% prevalence of resistance threshold. As drug A resistance hits this threshold, drug B is quickly phased in from 0% use to 100% use with a midpoint 0.5 years beyond the point at which drug A is lost. B) The probability of receiving each drug is updated over time. Drug B is phased in from a small initial uptake at time 0 (1.7%) to 50% use with a midpoint of 4 years. Complete random 50-50 allocation is reached within 9.5 years.

**Supplementary Figure 2.**
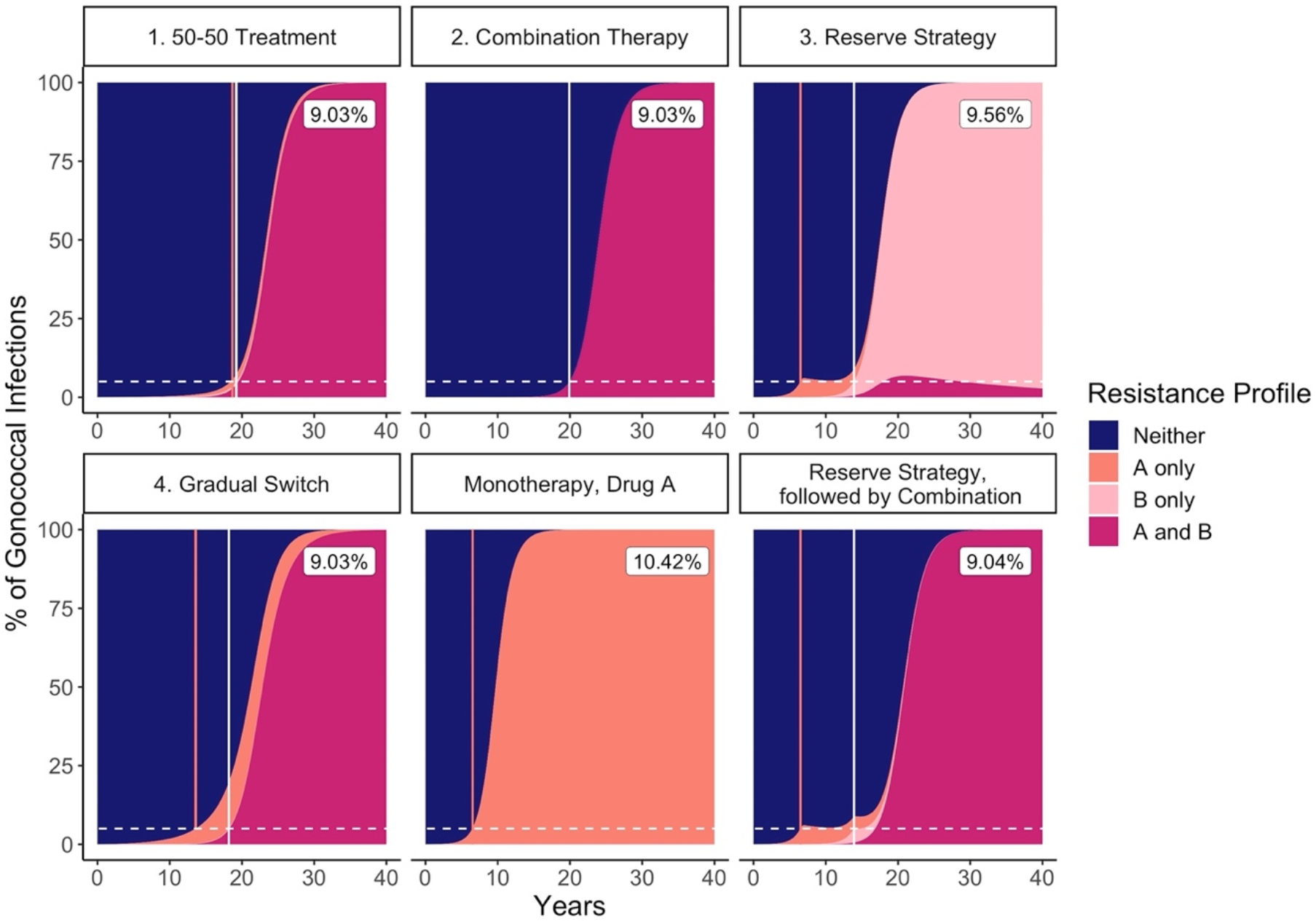
Percentage of gonococcal infections with each resistance profile over time, by strategy. The four antibiotic allocation strategies of interest are visualized along with i) continuing monotherapy using only drug A and ii) deploying combination therapy after each of drugs A and B are lost under the reserve strategy, for comparison. Dotted horizontal white lines indicate the 5% prevalence of resistance threshold, which warrants changes in treatment protocols per WHO guidelines. Solid vertical lines delineate the time at which only drug A is lost (salmon) and both drugs are lost (T_L_; white) by strategy. The population-level prevalence of gonococcal infection that the model re-equilibrates to is labeled in the upper right corner for each strategy.

**Supplementary Figure 3.**
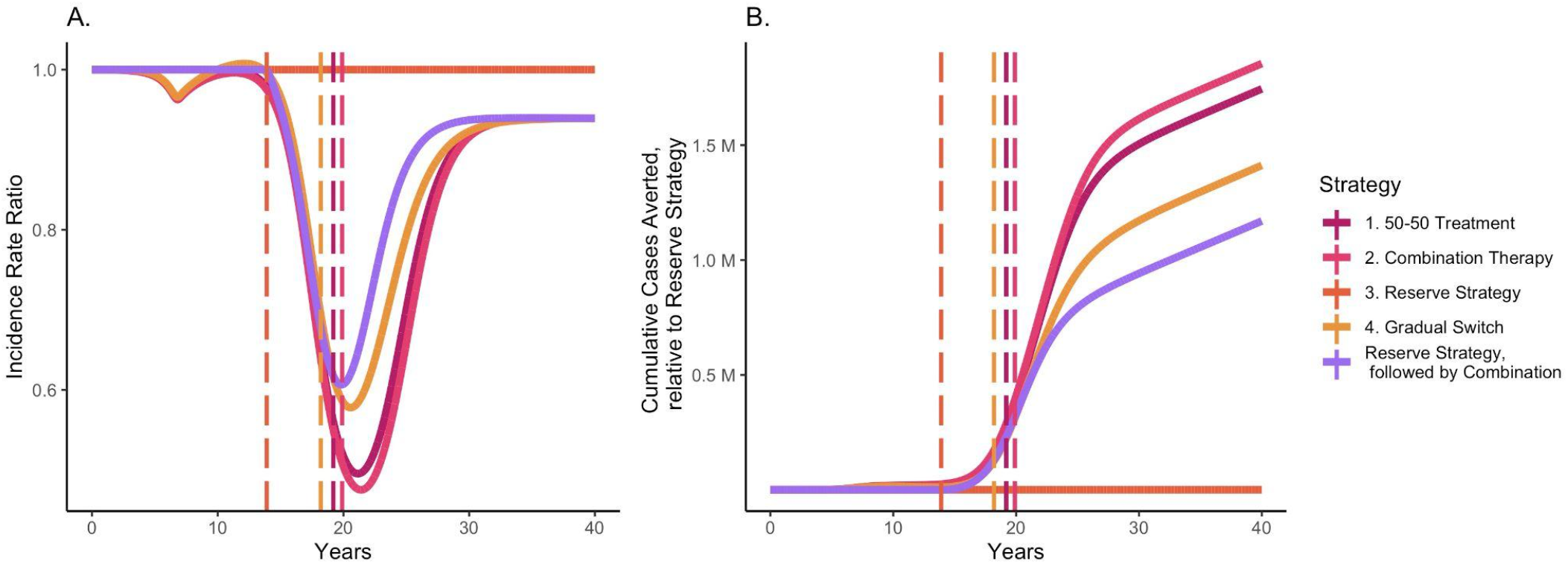
Dynamics of A) incidence rate ratios and B) cumulative number of incident infections averted over time, relative to the reserve strategy. Dashed vertical lines represent the end of the drugs’ lifespan (T_L_) for each introduction strategy under baseline model assumptions.

**Supplementary Figure 4.**
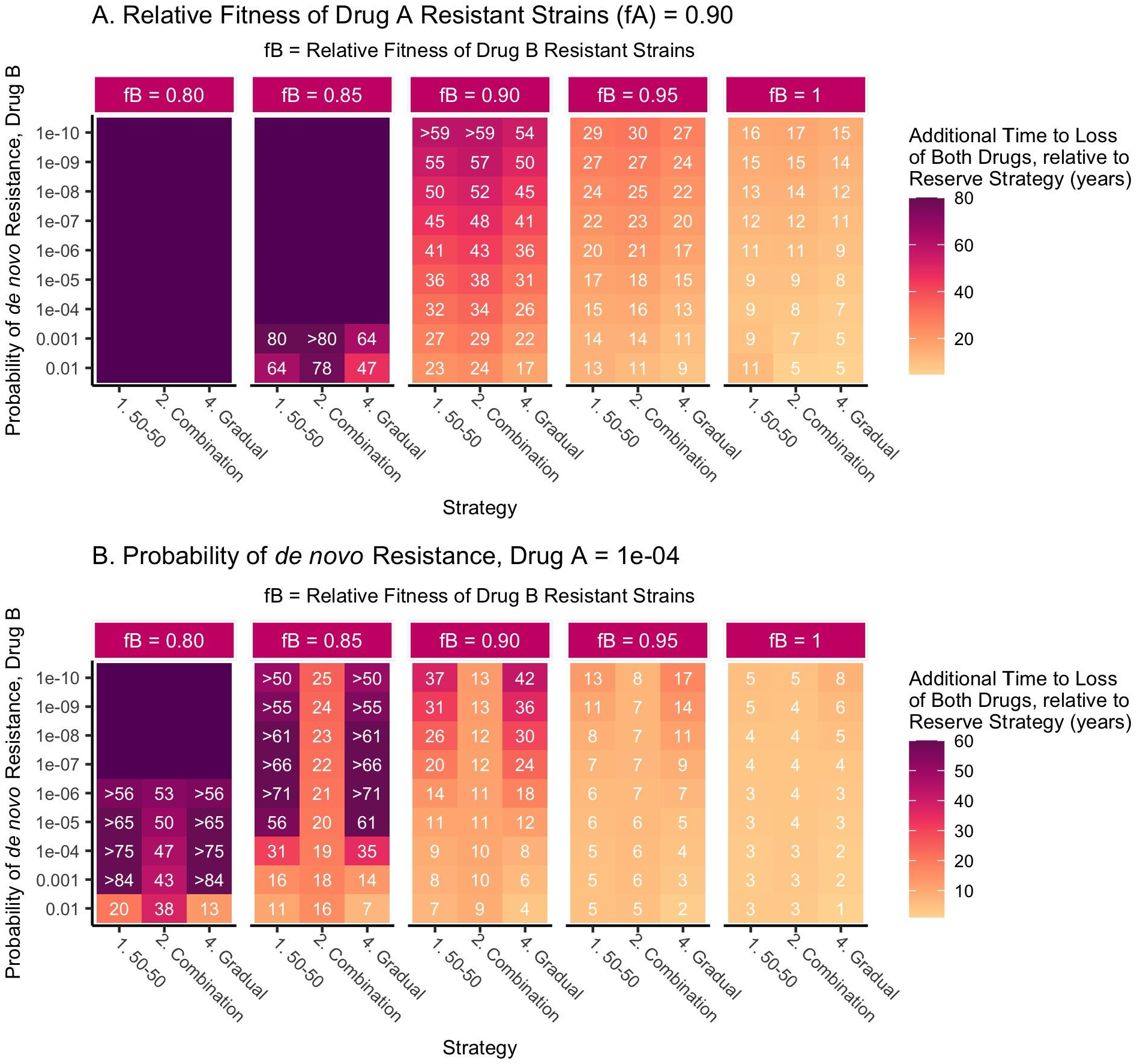
Additional time (in years) to loss of each of antibiotics A and B by strategy relative to that of the reserve strategy (T_L_|strategy - T_L_|reserve), based on properties of a new antibiotic B. Antibiotic A = ceftriaxone-like, Antibiotic B = new antibiotic. Strategies (x-axis) are compared over a range of plausible values for the probability of *de novo* resistance (or emergence of resistance upon treatment) with drug B (ω_B_; y-axis) and the relative fitness of drug B resistant strains (f_B_; vertical facets). **A)** The fitness cost associated with resistance to drug A is increased to 0.10. **B)** The probability of resistance upon treatment with drug A is increased to 10^−4^. If the lifespan of these drugs extended beyond the 100 years over which the model was run, that strategy’s results are shown either in relative terms for comparison or with an unlabeled dark purple tile, if no strategies on the x-axis had a defined T_L_ under that parameter set for drug B. Abbreviations: fB = f_B_ = fitness of drug B resistant strains relative to susceptible bacteria, fA = f_A_ = fitness of drug A resistant strains relative to susceptible bacteria, T_L_ = time in years until both drugs A and B have hit their 5% resistance thresholds, warranting new treatment recommendations.

### Technical Supplement

#### Model Structure

We use an adapted version of the single sex compartmental gonorrhea transmission model developed by Tuite et al.^1^ to describe our two-drug system. A visual overview of the model is presented in the main text (**Figure 1**). As previously described^1^, the model population is stratified into three sexual activity groups (k; low, intermediate, and high), characterized by annual rates of partner change. The model can be characterized as a susceptible-infectious-susceptible (SIS) model, where susceptible individuals (S) become infected and can then recover spontaneously or through antibiotic treatment. For those that seek treatment, one can receive drug A (ceftriaxone-like) and/or drug B (new antibiotic). Infections (I) are stratified by symptomatic (*Y*) versus asymptomatic (*Z*) infection, as well as by resistance profile, where each infection can be caused by bacteria resistant to drug A (ceftriaxone-like), drug B (novel antibiotic), neither, or both. The total size of the population (*N*) is set at 10^6^ and the absolute size of each sexual activity group at *N*_*k*_, with the relative size of each group fixed at *n*_*k*_.

Therefore,

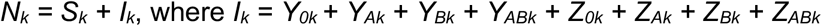

The relative rate of partner change (*rp*_*k*_) for each sexual activity group is drawn from previous estimates by Tuite et al.^1^ determined by data from the National HIV Behavioral Surveillance System^2^. The rate of partner change (*c*_*min*_) in the low activity group is estimated through the maximum likelihood estimation model fitting procedure. The annual rate of partner change for each activity group (*θ*_*k*_) is therefore described by the equation:

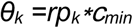

Assortativity between sexual activity groups is characterized by mixing parameter *ε* [which can range from 0 (random mixing) and 1 (fully assortative mixing) and is determined through model fitting]. This leads the probability of an individual from sexual activity group *i* coming into sexual contact with an individual from group *j* to be:

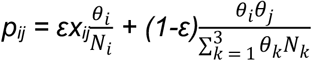

where *x*_*ij*_ is equal to 1 if *i = j* and equal to 0 if *‘ J K*. The rate of infection for susceptible individuals in sexual activity group *i* from infected partners of group *j* (*β*_*i←j*_) is proportional to this per capita probability of sexual contact between groups *i* and *j* (*p*_*ij*_), as well as the transmission probability per partnership (*b*). This relationship is defined by:

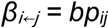

Therefore, the per capita transmission matrix ***β*** is a *k* ×*k* matrix with elements *β*_*i←j*_ at row i and column j, defined as the per capita rate of gonorrhea transmission to group i from group j. Note that the transmission matrix at element *β*_*i←j*_ is divided through by the size of group j’s population, which stays constant over time. As area is also held constant, and deemed not influential in per capita transmission rates, the model assumes constant density and yields results identical to frequency-dependent transmission.

#### Model Recovery from Gonococcal Infection

Parameters describing treatment and retreatment rates by infection type (symptomatic vs. asymptomatic), as well as the rate of natural clearance of infection, are presented in the main text (Table 1). These parameters can be used to calculate the average duration of an infection (years), which is (1/T_s_) for treated symptomatic infections, (1/T_m_) for treated asymptomatic infections detected via screening, and (1/δ) for infections that clear naturally. The treatment rate T_sr_ for those with an initial treatment failure assumes an average duration of infection three times that of those with initial treatment success; 1/T_sr_ represents the average time in years until successful retreatment, including the time it takes for: 1) the individual to receive the initial failed treatment, 2) the individual to re-seek care, and 3) the provider to identify and prescribe the correct antibiotic for retreatment. It is possible for resistance acquired upon initial treatment of a susceptible infection to then be retreated insufficiently with the same antibiotic before receiving a successful retreatment; in these instances, the average duration of infection is (1/T_s_ + 1/T_sr_), in years.

#### Model Equations

In matrix form, the model is described by the following system of differential equations, where ∘ denotes element-wise multiplication. All parameters are defined in the main text (**Table 1**). Bolded letters represent *k* ×*1* column vectors with compartmental variables for each sexual activity group (e.g., **S** = [S_1_,…S_k_]^T^) for *k* groups), with the exception of the *k* ×*k* matrix ***β***.

Strategies 1,3,4: Random (50-50) allocation, reserve strategy, and gradual introduction:

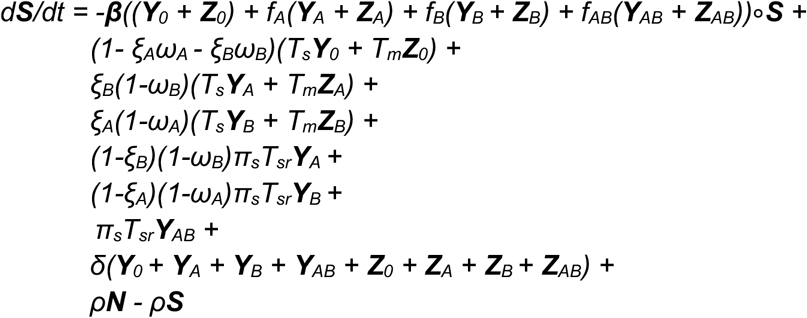

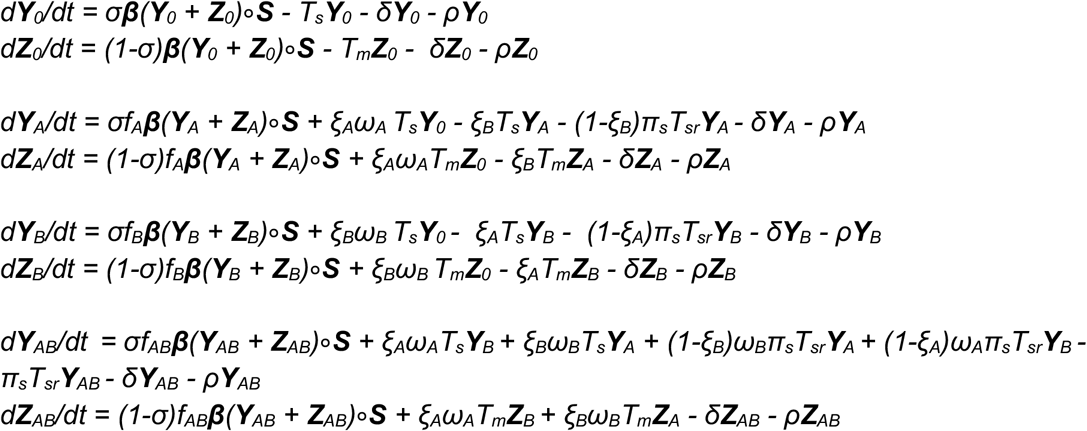

Strategy 2: Combination treatment:

Note: *ξ*_A_ and *ξ*_B_ are omitted from the following equations since both = 1.

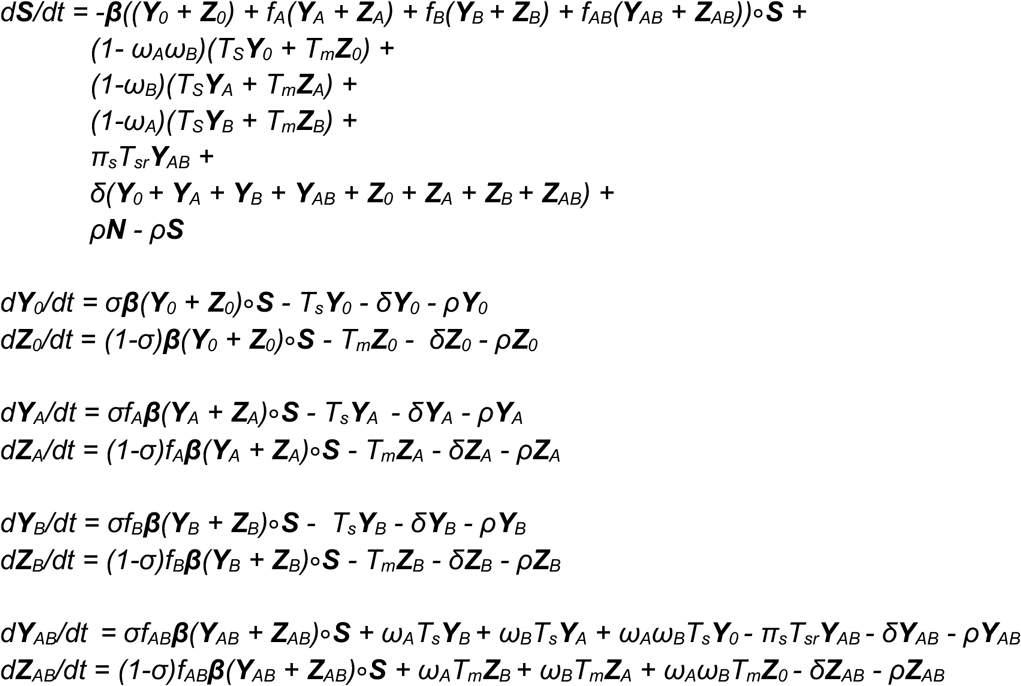

We calculate the number of incident infections at time *t*, or the overall force of infection across sexual activity groups, as λ_t_ = ***β***((***Y***_*0t*_ *+* ***Z***_*0t*_*) + f*_*A*_(***Y***_*At*_ *+* ***Z***_*At*_*) + f*_*B*_(***Y***_*Bt*_ *+* ***Z***_*Bt*_*) + f*_*AB*_(***Y***_*ABt*_ *+* ***Z***_*ABt*_))∘***S***_*t*_.

The overall prevalence of infection at time t can be calculated as Prev_t_ = (***Y***_*0t*_ *+* ***Z***_*0t*_ *+* ***Y***_*At*_ *+* ***Z***_*At*_ *+* ***Y***_*Bt*_ *+* ***Z***_*Bt*_ *+* ***Y***_*ABt*_ *+* ***Z***_*ABt*_*)/N*.

## References

1. Centers for Disease Control and Prevention. Sexually Transmitted Infections Treatment Guidelines, 2021 [Internet]. [cited 2022 Jul 13]. Available from: https://www.cdc.gov/std/treatment-guidelines/gonorrhea-adults.htm

2. Taylor SN, Marrazzo J, Batteiger BE, Hook EW, Seña AC, Long J, et al. Single-Dose Zoliflodacin (ETX0914) for Treatment of Urogenital Gonorrhea. N Engl J Med. 2018 Nov 8;379(19):1835–45.

3. Taylor SN, Morris DH, Avery AK, Workowski KA, Batteiger BE, Tiffany CA, et al. Gepotidacin for the Treatment of Uncomplicated Urogenital Gonorrhea: A Phase 2, Randomized, Dose-Ranging, Single-Oral Dose Evaluation. Clin Infect Dis Off Publ Infect Dis Soc Am. 2018 Aug 1;67(4):504–12.

4. Bonhoeffer S, Lipsitch M, Levin BR. Evaluating treatment protocols to prevent antibiotic resistance. Proc Natl Acad Sci U S A. 1997 Oct 28;94(22):12106–11.

5. Bergstrom CT, Lo M, Lipsitch M. Ecological theory suggests that antimicrobial cycling will not reduce antimicrobial resistance in hospitals. Proc Natl Acad Sci. 2004 Sep 7;101(36):13285–90.

6. Wang YC, Lipsitch M. Upgrading antibiotic use within a class: Tradeoff between resistance and treatment success. Proc Natl Acad Sci. 2006 Jun 20;103(25):9655–60.

7. Haber M, Levin BR, Kramarz P. Antibiotic control of antibiotic resistance in hospitals: a simulation study. BMC Infect Dis. 2010 Aug 25;10(1):254.

8. Joyner ML, Manning CC, Canter BN. Modeling the effects of introducing a new antibiotic in a hospital setting: A case study. Math Biosci Eng MBE. 2012 Jul;9(3):601–25.

9. Obolski U, Stein GY, Hadany L. Antibiotic Restriction Might Facilitate the Emergence of Multi-drug Resistance. PLOS Comput Biol. 2015 Jun 25;11(6):e1004340.

10. Tepekule B, Uecker H, Derungs I, Frenoy A, Bonhoeffer S. Modeling antibiotic treatment in hospitals: A systematic approach shows benefits of combination therapy over cycling, mixing, and mono-drug therapies. PLOS Comput Biol. 2017 Sep 15;13(9):e1005745.

11. Xiridou M, Soetens LC, Koedijk FDH, Sande M a. Bvd, Wallinga J. Public health measures to control the spread of antimicrobial resistance in Neisseria gonorrhoeae in men who have sex with men. Epidemiol Infect. 2015 Jun;143(8):1575–84.

12. Centers for Disease Control and Prevention. Sexually Transmitted Disease Surveillance 2020: Gonococcal Isolate Surveillance Project (GISP) Profiles [Internet]. [cited 2022 Jul 14]. Available from: https://www.cdc.gov/std/statistics/gisp-profiles/default.htm

13. Centers for Disease Control and Prevention. Sexually Transmitted Disease Surveillance 2020 [Internet]. [cited 2022 Jul 13]. Available from: https://www.cdc.gov/std/statistics/2020/overview.htm

14. Centers for Disease Control and Prevention (CDC). Increases in fluoroquinolone-resistant Neisseria gonorrhoeae among men who have sex with men--United States, 2003, and revised recommendations for gonorrhea treatment, 2004. MMWR Morb Mortal Wkly Rep. 2004 Apr 30;53(16):335–8.

15. Fingerhuth SM, Bonhoeffer S, Low N, Althaus CL. Antibiotic-Resistant Neisseria gonorrhoeae Spread Faster with More Treatment, Not More Sexual Partners. PLOS Pathog. 2016 May 19;12(5):e1005611.

16. Fingerhuth SM, Low N, Bonhoeffer S, Althaus CL. Detection of antibiotic resistance is essential for gonorrhoea point-of-care testing: a mathematical modelling study. BMC Med. 2017 Jul 26;15(1):142.

17. Tuite AR, Gift TL, Chesson HW, Hsu K, Salomon JA, Grad YH. Impact of Rapid Susceptibility Testing and Antibiotic Selection Strategy on the Emergence and Spread of Antibiotic Resistance in Gonorrhea. J Infect Dis. 2017 Nov 27;216(9):1141–9.

18. Johnson Jones ML, Chapin-Bardales J, Bizune D, Papp JR, Phillips C, Kirkcaldy RD, et al. Extragenital Chlamydia and Gonorrhea Among Community Venue-Attending Men Who Have Sex with Men - Five Cities, United States, 2017. MMWR Morb Mortal Wkly Rep. 2019 Apr 12;68(14):321–5.

19. Grov C, Cain D, Rendina HJ, Ventuneac A, Parsons JT. Characteristics Associated With Urethral and Rectal Gonorrhea and Chlamydia Diagnoses in a US National Sample of Gay and Bisexual Men: Results From the One Thousand Strong Panel. Sex Transm Dis. 2016 Mar;43(3):165–71.

20. Bolker B, Giné-sVázquez I, R Development Core Team. bblme: Tools for General Maximum Likelihood Estimation [Internet]. 2022. Available from: https://cran.r-project.org/package=bbmle

21. Jacobsson S, Golparian D, Oxelbark J, Franceschi F, Brown D, Louie A, et al. Pharmacodynamic Evaluation of Zoliflodacin Treatment of Neisseria gonorrhoeae Strains With Amino Acid Substitutions in the Zoliflodacin Target GyrB Using a Dynamic Hollow Fiber Infection Model. Front Pharmacol [Internet]. 2022 [cited 2022 Sep 28];13. Available from: https://www.frontiersin.org/articles/10.3389/fphar.2022.874176

22. Tuite AR, Rönn MM, Wolf EE, Gift TL, Chesson HW, Berruti A, et al. Estimated Impact of Screening on Gonorrhea Epidemiology in the United States: Insights From a Mathematical Model. Sex Transm Dis. 2018 Nov;45(11):713–22.

23. Vegvari C, Grad YH, White PJ, Didelot X, Whittles LK, Scangarella-Oman NE, et al. Using rapid point-of-care tests to inform antibiotic choice to mitigate drug resistance in gonorrhoea. Eurosurveillance. 2020 Oct 29;25(43):1900210.

24. Hui BB, Wilson DP, Ward JS, Guy RJ, Kaldor JM, Law MG, et al. The potential impact of new generation molecular point-of-care tests on gonorrhoea and chlamydia in a setting of high endemic prevalence. Sex Health. 2013 Aug;10(4):348–56.

25. Tapsall J, Anti-Infective Drug Resistance Surveillance and Containment Team. Antimicrobial resistance in Neisseria gonorrhoeae [Internet]. World Health Organization; 2001 [cited 2022 Jul 14]. Report No.: WHO/CDS/CSR/DRS/2001.3. Available from: https://apps.who.int/iris/handle/10665/66963

26. Soetaert K, Petzoldt T, Setzer R. Solving Differential Equations in R: Package deSolve. J Stat Softw. 2010;33(9):1–25.

27. Olesen SW, Grad YH. Deciphering the Impact of Bystander Selection for Antibiotic Resistance in Neisseria gonorrhoeae. J Infect Dis. 2020 Apr 1;221(7):1033–5.

28. Adamson PC, Lin EY, Ha SM, Klausner JD. Using a public database of Neisseria gonorrhoeae genomes to detect mutations associated with zoliflodacin resistance. J Antimicrob Chemother. 2021 Oct 11;76(11):2847–9.

29. Scangarella-Oman NE, Hossain M, Dixon PB, Ingraham K, Min S, Tiffany CA, et al. Microbiological Analysis from a Phase 2 Randomized Study in Adults Evaluating Single Oral Doses of Gepotidacin in the Treatment of Uncomplicated Urogenital Gonorrhea Caused by Neisseria gonorrhoeae. Antimicrob Agents Chemother. 2018 Nov 26;62(12):e01221–18.

## References

1. Tuite AR, Gift TL, Chesson HW, Hsu K, Salomon JA, Grad YH. Impact of Rapid Susceptibility Testing and Antibiotic Selection Strategy on the Emergence and Spread of Antibiotic Resistance in Gonorrhea. J Infect Dis. 2017;216(9):1141–1149. doi:10.1093/infdis/jix450

2. Centers for Disease Control and Prevention. HIV Infection Risk, Prevention, and Testing Behaviors among Men Who Have Sex With Men -- National HIV Behavioral Surveillance, 20 U.S. Cities, 2014. HIV Surveillance Special Report 15. Published January 2016. Accessed September 8, 2022. http://www.cdc.gov/hiv/library/reports/surveillance/#panel2

